# Evaluations of serological test in the diagnosis of 2019 novel coronavirus (SARS-CoV-2) infections during the COVID-19 outbreak

**DOI:** 10.1101/2020.03.27.20045153

**Authors:** Dachuan Lin, Lei Liu, Mingxia Zhang, Yunlong Hu, Qianting Yang, Jiubiao Guo, Youchao Dai, Yuzhong Xu, Yi Cai, Xinchun Chen, Kaisong Huang, Zheng Zhang

**Affiliations:** Guangdong Provincial Key Laboratory of Regional Immunity and Disease, Department of Pathology Biology, School of Medicine, Shenzhen University, Shenzhen, China; National clinical research center for infectious diseases, Guangdong Key Lab for Diagnosis &Treatment of Emerging Infectious Diseases, Shenzhen Third People’s Hospital, Southern University of Science and Technology, Shenzhen, China; Department of clinical laboratory, Shenzhen Baoan people’s hospital, The Second Affiliated Hospital of Shenzhen University, Shenzhen, China

**Author notes:** These authors contributed equally to this work (Dachuan Lin, Lei Liu).

**Keywords:** SARS-CoV-2, serological testing, chemiluminescence immunoassay, IgM and IgG

## Abstract

The ongoing SARS-CoV-2 outbreak has killed over twenty-one thousand and sickened over four hundred thousand people worldwide, posing a great challenge to global public health. A sensitive and accurate diagnosis method will substantially help to control disease expansion. Here, we developed a chemiluminescence-immunoassay method based on the recombinant nucleocapsid antigen and the magnetic beads for diagnosis of SARS-CoV-2 infections and surveillance of antibody changing pattern.

Serums from 29 healthy individuals, 51 tuberculosis patients, and 79 SARS-CoV-2 confirmed patients were employed to evaluate the performance of this approach. Compared to the IgM testing, the IgG testing was more reliable in which it identified 65 SARS-CoV-2 infections from the 79 confirmed patients and only two false-positive cases from the 80 control group with a sensitivity and specificity reaching 82.28% and 97.5%, respectively. However, only a slight difference (not statistically significant) in the detected cases of SARS-CoV-2 infections was observed between the IgM and IgG testing manner in patients at a different time of onset of disease. A performance comparison between an ELISA kit using the same nucleocapsid antigen and our chemiluminescence method was undertaken. The same false-positive cases were seen in both methods from the paired control group, while ELISA kit can only detect half of the SARS-CoV-2 infections from paired SARS-CoV-2 confirmed patients group than that of the chemiluminescence method, indicating a higher performance for the chemiluminescence-immunoassay approach. Together, our studies provide a useful and valuable serological testing tool for the diagnosis of SARS-CoV-2 infections in the community.

## Introduction

*Coronavirus*, belonging to the family of *Coronavirdiae* and order of *Nidovirales*, is a group of enveloped, non-segmented positive-sense RNA virus that has been reported to be able to infect humans and a wide range of animals including cattle, swine, chicken, cat, horse, camels, rodent, bats and snakes and so forth (1-3). Based on the genetic properties, coronavirus was further divided into four genera, namely *Alphacoronavirus, Betacoronavirus, Gammacoronavirus*, and *Deltacoronavirus (4)*. Prior to December 2019, a total of six coronaviruses have been documented to be capable of causing disease in humans. These include two strains from *Alphacoronavirus* (HCoV-229E and HKU-NL63) and four from *Betacoronavirus* subfamily (HCoV-OC43, HCoV-HKU1, SARS-CoV and MERS-CoV) (5-10). Among them, the Severe Acute Respiratory Syndrome Coronavirus (SARS-CoV) and Middle East Respiratory Syndrome coronavirus (MERS-CoV) are the most well-described as they directly led to two deadly large-scale outbreaks globally, with 8,096 cases infections and roughly 10 percent mortality and 2,494 cases and 34.4 percent mortality, respectively(9, 10).

Recently, the outbreak of a severe pneumonia COVID-19 was confirmed to be caused by the 2019 novel coronavirus infections (SARS-CoV-2) that was originated from a seafood wholesale market in Wuhan, China(11). So far, this novel coronavirus has spread throughout the whole of China and over 198 countries globally, causing over468,905 laboratory-confirmed cases of infections with 21,200 people dead posing a great threat to the global public health (http://2019ncov.chinacdc.cn/2019-nCov/). Besides, there are still numerous suspected cases and a myriad of medical monitoring people who were quarantined in specialized hospitals or at homes because of their previous epidemiological link to confirmed SARS-CoV-2 patients. All of these put an extreme burden on the emergency, hospital and public health system particularly the epidemic zone worldwide. Therefore, a timely, sensitive and accurate diagnosis approach is urgently needed and of pivotal importance for surveillances of disease dissemination and the prevention of further expansions. Conventional diagnosis methods such as virus culture and microscopic analysis are generally time-consuming and labor-intensive with limited sensitivity (12, 13). In contrast, the last decade emerged molecular biologic and serologic approaches, such as TaqMan Real-Time Reverse Transcription Polymerase Chain Reaction (RT-PCR), Enzyme-linked immunosorbent assay, colloidal gold immunochromatography and direct chemiluminescence immunoassay(CLIA), can be developed into a rapid and effective tool for detections of respiratory pathogens infections, even though in certain circumstances molecular biologic method like RT-PCR had a low sensitivity for specimens from upper respiratory tract (14-17).

In this study, we developed a chemiluminescence immunoassay method to specifically detect the induced antibody IgM and IgG by SARS-CoV-2 using the recombinant nucleocapsid (YP_009724397.2) and evaluate its sensitivity and specificity in detections of SARS-CoV-2 infected patients. High sensitivity and specificity results indicate this chemiluminescence immunoassay method in combination with RT-PCR method can serve as highly sensitive and accurate tools for the diagnosis and screen of SARS-CoV-2 infections in the community.

## Material and methods

### Participants and specimens

In Shenzhen city, China, patients infected by SARS-CoV-2 were all eventually admitted into a specialized hospital (the third people’s hospital of Shenzhen) for quarantines and treatments. In this study, a total of 29 healthy individuals, 51 tuberculosis patients and 79 SARS-CoV-2 patients were enrolled for serological testing. Twenty-nine healthy people were recruited from the Shenzhen University aging from 16 to 72 years old. Fifty-one tuberculosis patients were enrolled from the Shenzhen Baoan hospital, and their mycobacterium tuberculosis infections were previously confirmed by sputum smear acid-fast-bacilli analysis, chest radiograph and the QuantiFERON®–TB Gold test.

COVID-19 patients were randomly enrolled from the third people’s hospital of Shenzhen, and their SARS-CoV-2 infections were confirmed by combinations of epidemiological risk, clinical features and positive detections of SARS-CoV-2 RNA in respiratory specimens using the National Medical Production Administration authorized GeneoDX kit according to the official instruction for diagnosis and treatment of 2019 novel coronavirus infections issued by the National Health Commission of the People’s Republic of China. All healthy cohorts and tuberculosis patients had no epidemiological risks, and they were persistently negative for SARS-CoV-2 RNA detections in at least three respiratory specimens’ tests using above mentioned GeneoDX kit. Peripheral blood samples were collected into EDTA and sodium heparin containing tubes, and then the serum was separated by centrifugations (800g ×10 minutes) for immediate testing or stored at −80°C until used. Verbal Informed consent was obtained from all individual participants.

### SARS-CoV-2 Nucleic acid

Total nucleic acid for collected respiratory specimens was extracted in BSL-3 laboratory using the nucleic acid extraction and purification kit from Huayin Bio-Tech (Shenzhen, China) according to the manufacturer protocol. Detections of SARS-CoV-2 RNA were performed using the National Medical Production Administration authorized GeneoDX kit (Taqman RT-PCR method, targeting the ORF1ab and N genes) according to the manufacturer instructions.

### Development of chemiluminescence immunoassay and test procedures

After the transcription of extracted SARS-CoV-2 genome RNA into the cDNA, the coding regions (YP_009724397.2) were then amplified and cloned into the pET30a vector. The recombinant full-length nucleocapsid antigen was expressed in engineering *E*. coli BL21 (DE3) strains and purified using the Ni-NTA resin (Darui Biotech, China). Magnetic beads Magnosphere™ MS300 used in this study are commercially available in the JSR Corporation, Tokyo, Japan. Recombinant nucleocapsid antigens were coupled to these tosyl magnetic beads using the catalytic reagent solution (3M Ammonium sulfate / 0.1M Borate buffer, pH9.5) according to the manufacture’s instruction, and the resultant beads were further blocked by 0.05% BSA for six hours at 37 °C. The following testing and detection procedure was automated on a chemical immuno-luminescence analyzer ACCRE6 (Tianshen Tech, Shenzhen, China). It was comprised of those following steps. 50 microliter pure serum was firstly incubated with the magnetic beads that were coupled with antigens for 5 minutes at 37 °C. Subsequently, the unbound substance was gently removed and then washed by Tris buffer for three times. Alkaline phosphatase labeled anti-human immunoglobulin (50µg/ml) was added and further incubated for 5 minutes at 37 °C in the Mes Buffer. After three times washing to remove unbound materials, the lumigen APS-5 substrate (50ug/ml) was eventually added. Ultimately, the light signal was measured by the photomultiplier in ACCRE6 (Tianshen Tech, Shenzhen, China) as relative light units, and the whole testing can be completed in 23 minutes. Confirmed SARS-CoV-2 positive-serum and negative-serum were used as controls in each set test.

### Detections of IgG and IgM by a commercial ELISA kit

In parallel testing, the commercial enzyme-linked immunosorbent assay kit (Darui Biotech, CHINA) for detections of the anti-SARS-CoV-2 IgG and IgM antibody was used to measure the SARS-CoV-2 antibody level in above mentioned COVID-19 patients and control individuals. For the principle of this ELISA kit, the specific SARS-CoV-2 nucleocapsid protein and anti-human IgM monoclonal antibody were firstly coated on the plates, respectively. Subsequently, the 100 µl of 100-fold diluted serum was added and then incubated for 60 minutes at 37 °C. After five times washing by PBST buffer, the horseradish peroxidase (HRP) labeled mouse anti-human IgG antibody or HRP-labeled SARS-CoV-2 nucleocapsid antigen was added for 30 minutes incubation at 37 °C. Fifty microliter TMB substrate was then added for 15 minutes incubation after the second time washing by PBST buffer. The stopping solution was eventually added to suspend the reaction, and OD 450/630 values were immediately measured using the Varioskan LUX™ Multimode Microplate Reader. The cutoff values for positive were set based on the manufacturer’s recommendations.

### Ethical statement

The internal use of collected samples for diagnoses of etiological agents and serological research was approved by the Ethical Committee in the third people’s hospital of Shenzhen (SZTHEC2016001).

### Statistical analysis

All statistical analysis was performed in GraphPad Prism 7 software. The One-way ANOVA test was used to analyze the average RLU values difference between different participant groups. The chip-square and Fisher’s exact test was used for comparing the difference between the analyzed groups. The difference was considered significant when a p-value is < 0.05.

## Results

### Detections of IgG and IgM antibodies induced by SARS-CoV-2 in serum and overall specificity and sensitivity assessments

To assess the specificity and sensitivity of the chemiluminescence immunoassay method developed based on the recombinant nucleocapsid antigen, serum from 29 healthy individuals, 51 tuberculosis patients and 79 confirmed SARS-CoV-2 patients were employed and tested. More than six-fold and eight folds higher average RLU (relative light units) values were observed in the SARS-CoV-2 patients group in the IgM testing compared to that of the healthy cohort and tuberculosis patients (Figure 1A). This average RLU difference is more dramatic when it comes to the IgG testing reaching 60 and 70 fold increase in SARS-CoV-2 patients in comparison with the healthy and tuberculosis group, respectively (Figure 1B). A Receiver Operating Characteristic curve was then obtained based on these RLUs for the SARS-CoV-2 patients group and control group that consists of healthy cohort and tuberculosis patients. According to the ROC curve and analysis results, we recommend a cutoff setting for IgM (RLU 162296) and IgG (336697), in which the calculated sensitivity and specificity for IgM were 82.28% and 81.25%, and 82.28% and 97.5% for IgG, respectively.

**Figure 1.**
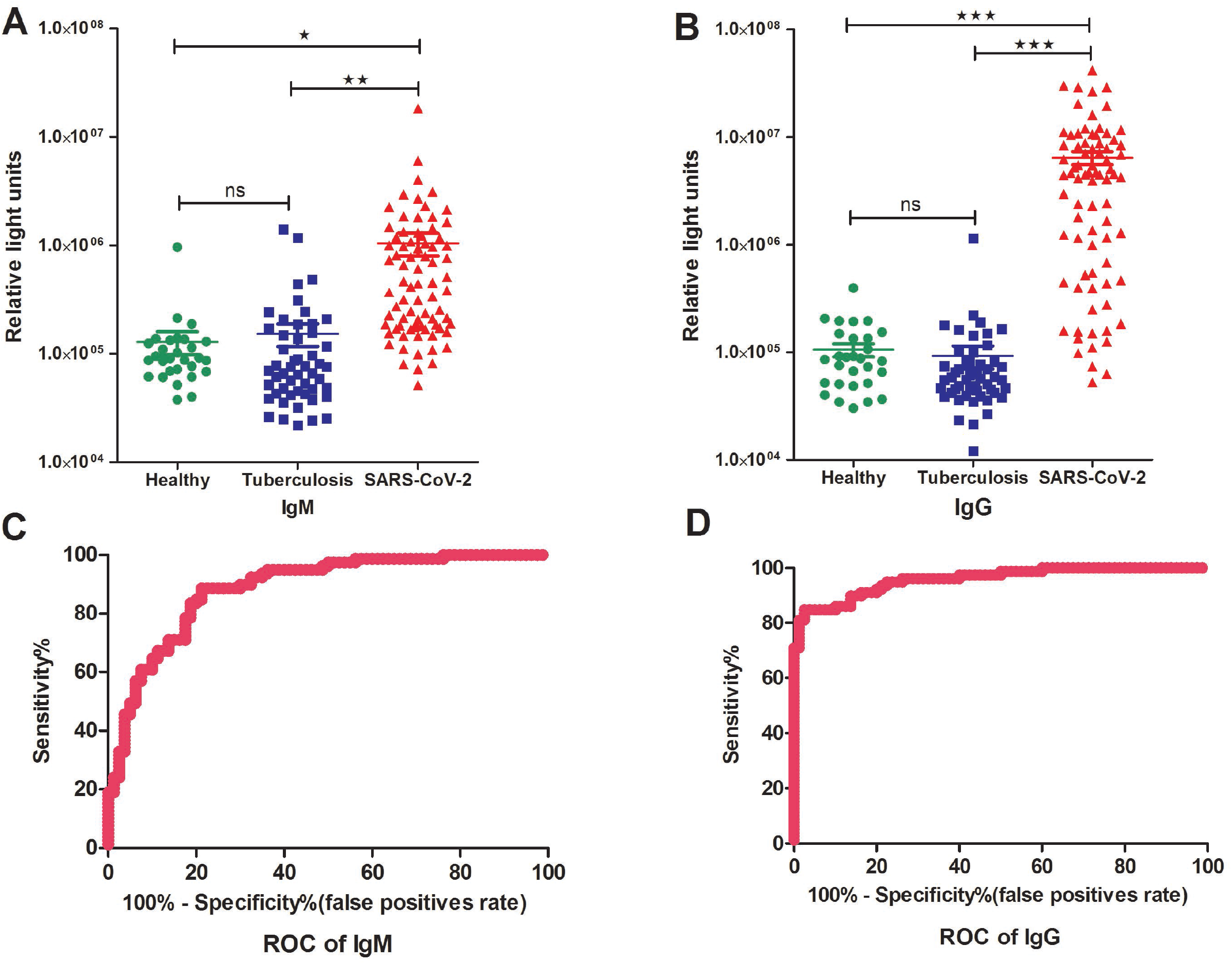
Detections and measurements of the SARS-CoV-2 IgM and IgG antibody in healthy people, tuberculosis patients and SARS-CoV-2 confirmed patients using the chemiluminescence immunoassay method (A and B). The average results were expressed as mean ± SEM of all individuals. Receiver Operating Characteristic curves for IgM (C) and IgG(D) were obtained based on the RLU for the SARS-CoV-2 patient group and the control group consisting of healthy cohorts and tuberculosis patients.

Based on this cutoff and using the IgM testing, we identified 15 cases and 65 cases as SARS-CoV-2 positive from the control group (80 cases) and the SARS-CoV-2 confirmed group (79 cases), respectively (Table 1). In contrast, using the IgG testing, we only detected two false-positive cases from the control group, which is in line with the higher specificity for IgG (97.5%) compared to that of the IgM testing (81.25%) as above described.

**Table 1.** Evaluations of a chemiluminescence immunoassay method for diagnosis of SARS-CoV-2 by detections of specific IgM and IgG in the patient’s serum.

| Participants category /total cases | IgM positive cases / (%) | IgG positive/ Cases / (%) | IgM or IgG positive cases/ (%) | IgM and IgG positive cases (%) |
| --- | --- | --- | --- | --- |
| Healthy cohorts and tuberculosis patients/80 cases | 15 /18.75% | 2 /2.50% | 16 /20.00% | 1 /1.25% |
| SARS-CoV-2 confirmed patients /79 cases | 65 /82.28% | 65 /82.28% | 72 /91.14% | 58/73.42% |

### The links between disease onset time, ages and IgM and IgG productions and detection efficiency

To explore whether the onset time was significantly linked with the detection sensitivity by this serological chemiluminescence method, comparison and statistical analysis of the sensitivity rates between different onset time patient categories was undertaken. No statistically significant difference was observed between the IgM and IgG testing results in the patients with the same onset time, although two more cases from 12 cases were detected by IgM testing compared to that of IgG testing in patients less than the one-week onset of disease (Table 2, p-value >0.05). In stark contrast, two more cases SARS-CoV-2 patients were identified by IgG testing than that of IgM testing in patients with more than two weeks onset of disease (p-value >0.05). In addition, we also compared the detection rates between the different age groups people, and we found that a significantly lower detection rate in both lgM and lgG testing manner for individual group younger than 18 years old was observed compared to that of people aging from 18 to 65 (p-value < 0.01) or over 65 years old (p-value >0.05) (Supplementary Table S1). No statistically significant differences were observed for male and female groups as well.

**Table 2.** Comparison of SARS-CoV-2 detections results in patients with different onset time between the IgM and IgG approach

| Days after onset | Total cases | IgM positive cases / (%) | IgG positive/ cases / (%) | IgM or IgG positive cases/ (%) | IgM and IgG positive cases (%) |
| --- | --- | --- | --- | --- | --- |
| 0-3 | 4 | 4/100% | 4/100% | 4/100% | 4/100% |
| 4-7 | 8 | 6/75% | 4/50% | 6/75% | 4/50% |
| 8-14 | 33 | 24/72.73% | 24/72.73% | 29/87.88% | 19/57.58% |
| >14 | 34 | 31/91.18% | 33/97.06% | 33/97.06% | 31/91.18% |

### Comparisons with other ELISA kit

To further characterize the patient’s immune response to the SARS-CoV-2 antigens and to prove the feasibility of the practical application of this serological testing kit in clinical diagnosis, 64 paired serum from the above-mentioned control cohorts and 65 COVID-19 patients were also examined using a recently developed commercial available ELISA Kit. A total of 14 false-positive cases (21%) were identified by IgM testing in both methods. A very lower false-positive rate was observed in IgG testing in both methods. Compared to the ELISA kit, a significantly higher detection rate for SARS-CoV-2 in both IgM and IgG testing manners was seen in our chemiluminescence method, suggesting a higher sensitivity of our approach compared to the tested ELISA kit (Table 3, p-value < 0.001).

**Table 3.** Detection differences between the chemiluminescence and ELISA method.

| Methods | Control group (total 64 cases) |  | SARS-CoV-2 confirmed patients (total 65 cases) |  |
| --- | --- | --- | --- | --- |
|  | IgM/false-positive /% | IgG/ false-positive /% | IgM positive /% | IgG positive /% |
| ELISA | 14 /21.8% | 0 / 0% | 30 / 46.1% | 15 / 23% |
| chemiluminescence | 14 /21.8% | 2 / 3.1% | 55 / 84.6% | 53 / 81.5% |
| Identified in both | 3 / 4.6% | 0 / 0% | 28 / 43% | 15 / 23% |

## Discussion

Compared with the conventional virological methods, the molecular biologic TaqMan RT-PCR method has been widely used for clinical diagnosis of respiratory pathogens infections particularly in the initial phase of disease because of its high specificity property (18, 19). Nevertheless, a relatively low sensitivity (30%-50%) in single upper respiratory specimen testing has been commonly reported including the well-appreciated methods for SARS-CoV detections (20). Furthermore, since the SARS-CoV-2 expansion from 2020 January, several cases reported that consecutive negative detections of SARS-CoV-2 RNA were observed for upper respiratory specimens testing like throat swabs in patients with apparent clinical symptoms, and the positive results can only be achieved by collecting the bronchoalveolar lavage fluid for re-testing. Hence, a sensitive serological diagnosis method can serve as a very useful compensation tool for current clinical diagnosis situations. Our results demonstrated that a single IgG testing is feasible in the clinical diagnosis for SARS-CoV-2, as a higher specificity and sensitivity were observed in our chemiluminescence method. In the humoral immune response, the antibody IgM was generally produced earlier than the IgG isotype as the IgM can be expressed without the isotype switching. Unexpectedly, we only observed a slight detection rate difference (not statistically significant) between these two antibody isotype testing manner in patients in the first week or more than two weeks after onset of disease. However, compared to the IgG approach, our IgM method showed a lower specificity (higher false-positive cases) in our testing. As the IgM and IgG using the same pure recombinant antigen and coupling condition (supplementary figure S1), the detection specificity difference is more likely linked to intrinsic antibody traits and concentration differences in the patients’ blood.

We noted that four patients with clinical symptoms less than four days were simultaneously detected by both IgM and IgG testing. A close examination of their medical record reveals that all of them had previous contact with confirmed SARS-CoV-2 individuals in at least 16 days ago, pointing to the possibility that they were probably asymptomatically infected by SARS-CoV-2 for certain time already. Fourteen cases (17.7%) from 79 SARS-CoV-2 confirmed patients were not identified by our serological testing method (both the IgM and IgG manner). Interestingly, of them, seven people were younger than 8-year-old or over 70-year-old. These people generally have low immunity in which a clinical symptom may occur rapidly upon exposure to the SARS-CoV-2, and we speculate that the antibodies in these people may not develop well yet when testing. More investigations are warranted to uncover the real situations. When comparing the detection rates in different age groups by our method, we noted that a significantly lower detection rate in both lgM and lgG testing manner for the individual group younger than 18 years old was observed compared to that of people aging from 18 to 65. An in-depth look at the days after onset for these 12 individuals younger than 18-years-old, the symptom onset time for all the 12 people are less than 14 days with six people even less seven days (Supplementary Table S2). The lower detection rate for these 12 people younger than 18-years-old was likely associated with no or less production of antibodies in them yet when we collected the serums.

In our parallel performance testing, the same antigen of nucleocapsid protein was used in both the commercially available ELISA kit and our chemiluminescence immunoassay. Unexpectedly, a significantly higher sensitivity was observed in our method compared to the ELISA kit. This sensitivity difference may be partially attributed to the difference in the serum amount for the first incubation step. On the other side, the intrinsic method difference including the aspect of binding surface interaction and mode of separations of the unbound material can also contribute to the sensitivity difference in the chemiluminescence immunoassay and ELISA method.

In conclusion, in this study, we developed and evaluated a serological chemiluminescence immunoassay testing technique for clinical diagnosis of SARS-CoV-2 infections using the recombinant nucleocapsid antigen. This high sensitivity and specificity chemiluminescence immunoassay method combined with the RT-PCR method can doubtless significantly improve the clinical diagnosis for SARS-CoV-2 and contribute to the control of COVID-19 expansion globally.

## Data Availability

All the data referred to in the manuscript has been included in the main text and supplementary files.

## Acknowledgment

We are very grateful to Prof. Yingsong Wu at Southern Medical University for his kind gifts of several proteins for our trail test and helpful suggestions for the whole project. This work is supported by Guangdong Provincial Science and Technology Program (No. 2019b030301009), the National Natural Science Funds of China (81802060), the start-up funding of Shenzhen University and the National Science and Technology Major Project (2017ZX10201301).

---

Supplementary figure S1. SDS-PAGE of the purified recombinant nucleocapsid antigen, M band indicates the marker, 1-3 band are the purified recombinant nucleocapsid protein.

